# Prevent the resurgence of infectious disease with asymptomatic carriers

**DOI:** 10.1101/2020.04.16.20067652

**Authors:** Zhechun Zhang

## Abstract

After reaching the peak of the COVID-19 outbreak, many regions are in search of a reopening plan that would minimize resurgence. Here we show to prevent resurgence local governments need to estimate how early and how many COVID-19 cases can be detected. It is safe to end social distancing if the majority of cases are detected early in the infection window. If cases are detected later, a 2-layer quarantine may be necessary to prevent resurgence. If fewer cases are detected, community events, schools, and large businesses may need to remain in virtual mode. Our results demonstrate that detecting more cases earlier is essential for preventing COVID-19 resurgence.

---

COVID-19 is an infectious disease with asymptomatic carriers^1,2^. An estimated 25%-43% of COVID-19 cases are asymptomatic^3,4^ (Fig. 1). In symptomatic cases, the disease can be transmitted 1-2 days before the onset of symptoms^1,2^ (Fig. 1). Asymptomatic transmission makes COVID-19 challenging to track and study. Unless everyone takes the test, only a portion of COVID-19 cases can be detected and quarantined (Fig. 1). Limited detection also makes estimating the reproduction number of COVID-19 more challenging than other infectious diseases such as SARS. How to prevent the resurgence of infectious disease with asymptomatic carriers is an open and urgent question.

**Figure 1.**
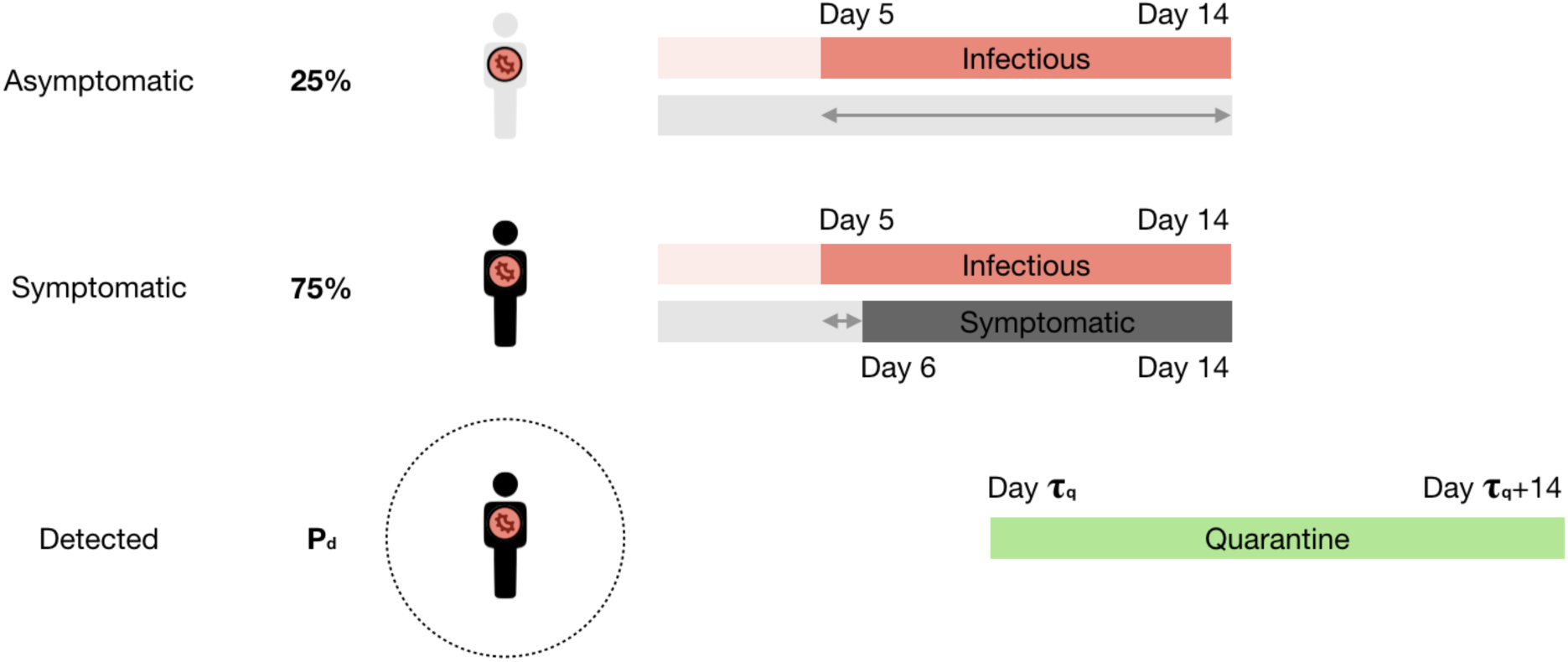
Asymptomatic transmission of COVID-19. Schematics showing the onset of COVID-19 transmission (red), symptom (grey), and quarantine (green). Arrows denote windows of asymptomatic transmission.

To address this question, we developed a microsimulation model^5,6^ where only a portion of cases is detected and quarantined. Disease carrier is infectious between days 5 and 14 post-infection^1,2^ (Fig. 1). Between days 6 and 14, some carriers are detected by testing (Fig. 1), resulting in the quarantine of confirmed carriers and their close contacts. We applied this model to a hypothetical region similar to Massachusetts, with 6.8 million residents, 3.9 million employees, and 2.5 million households (Fig. S1). Patient 0 was introduced on February 26 at a company conference; K-12 schools and colleges were closed on March 14; non-essential works were shut down on March 22 (Fig. S2).

We performed multiple tests to validate this model. First, we compared the number of cases in the simulation to the number of confirmed cases reported by the Massachusetts Department of Public Health (MDPH). Because the latter depends on testing availability and turnaround time, it sets a lower bound of actual cases approximately one week before the report date (Fig. S3). Second, we compared the number of deaths predicted by the simulation to the number of deaths reported by MDPH. Although the reported deaths is also subject to undercounting^7^, it provides an estimate of actual cases 1-2 weeks before the report date (Fig. S4). Here we assumed the death rate is ~1% as reported in Korea, where extensive testing started early in the outbreak. Third, our model predicts a peak date of reported cases on April 24 (Fig. S5). The prediction was made on April 15, posted on April 19^8^, and is consistent with the peak between April 23 and April 25 reported by MDPH (Fig. S6). The University of Washington model (covid19.healthdata.org accessed on April 15) predicted a peak date of resource use on April 28 (Fig. S7). Fourth, our model predicts a total death of 9,106 by August 5th, 11% more than the University of Washington model (Fig. S7). As of April 30, Massachusetts has seen 3,562 deaths, a figure that is likely an undercount^7^. Fifth, our model predicts a serial interval of 5.04 +/− 0.04 days, consistent with the reported interval of 4.4-7.5 days^1^. Last, our model predicts a reproduction number of 5.96 +/− 0.18. This number is higher than the COVID-19 estimate^1,9^ based on the early outbreak in Wuhan, where testing was limited. This is also higher than SARS^10,11^. Therefore, we modeled a disease that is more infectious than SARS.

According to the model, the reopening plan needs to be made based on estimated **P_d_**, the percentage of cases detected, and **τ_q_**, the time quarantine starts. The higher the **P_d_**, the lower the τ_q_, the less restrictive the reopening plan needs to be (Fig. 2). Social distancing can be lifted if 70% of cases can be detected, and quarantine starts within one week of infection (Fig. 2 and 3). These conditions are difficult, if not impossible, to satisfy. **P_d_** = 70% requires detection of most, if not all, symptomatic cases since asymptomatic cases account for 25% or more of total cases of COVID-19^3,4^. **τ_q_** = 7d requires quarantine to start within 1-2 days of the onset of symptoms.

**Figure 2.**
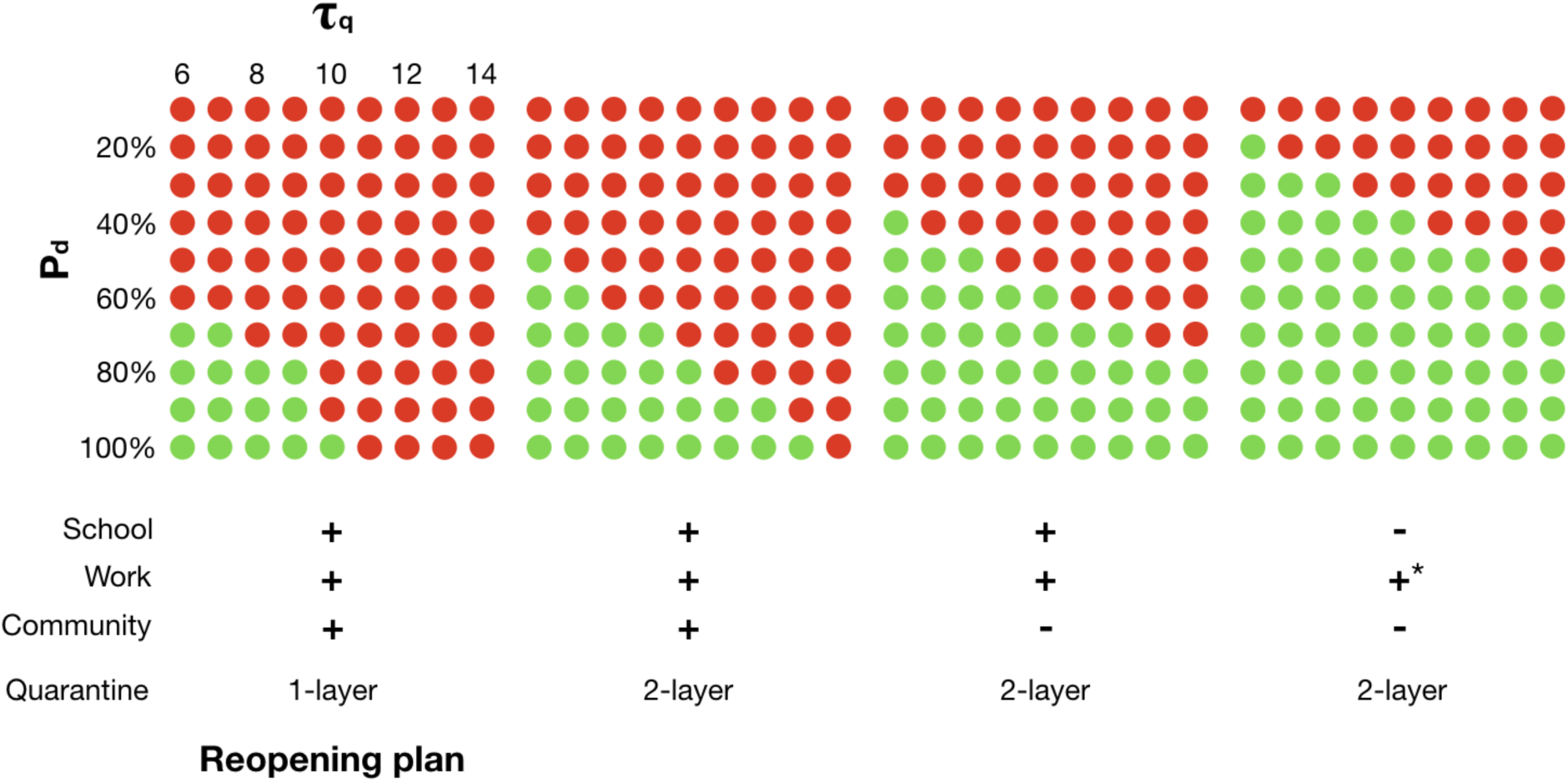
Resurgence as a function of P_d_, τ_q_, and reopening plan. Here **P_d_** is the percentage of cases detected, **τ_q_** is the time when quarantine starts. A red dot denotes resurgence after reopening.

**τ_q_** is an often neglected but critical variable. **τ_q_** determines whether 2-layer quarantine is necessary. If social distancing measures are lifted, but quarantine does not start within ten days of infection, a 100% detection rate is not sufficient to prevent resurgence (Fig. 2 and 3c). In general, the number of layers required is determined by two timescales: τ_q_, the time when quarantine starts, and τ_i_, the time when the patient becomes infectious. τ_i_, according to recent studies, is 4-5 days^1,2^. Currently, it can take another 4-5 days altogether for the test to come back positive and for public health officials to reach close contacts. At the time quarantine starts (τ_q_), the disease might have spread from the first layer of contacts to the next layer (Fig. 3a s).

**Figure 3.**
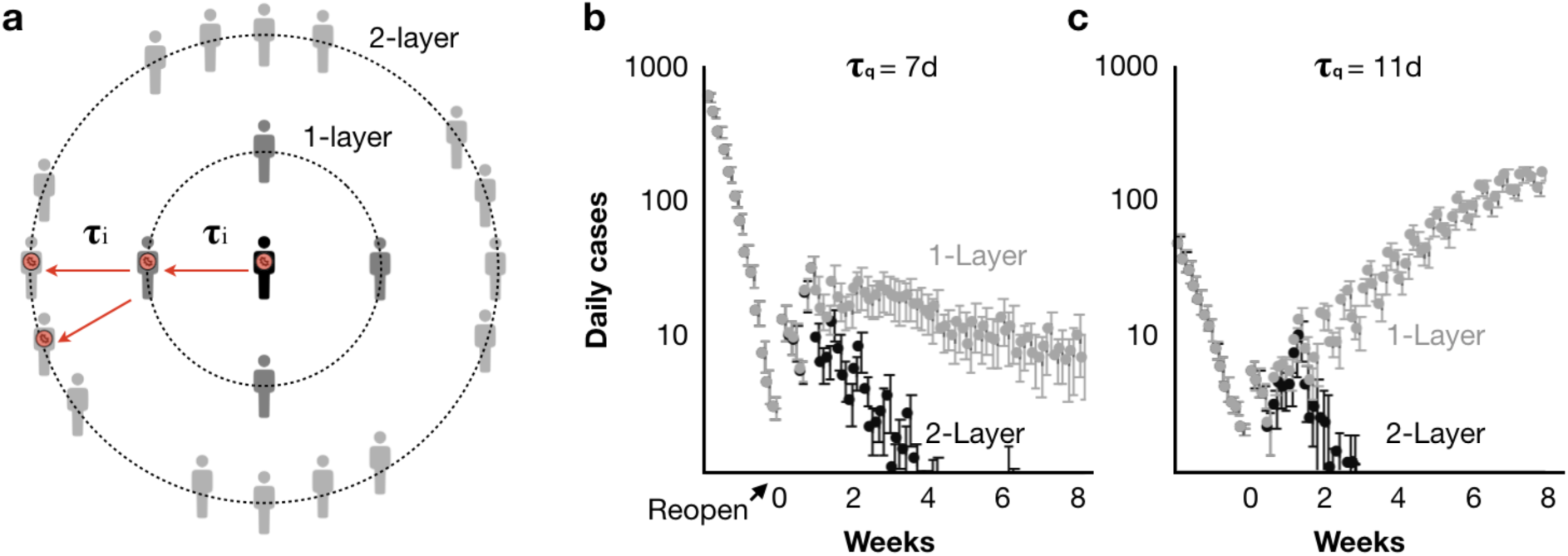
τ_q_ determines whether 2-layer quarantine is necessary. **a**, schematics of quarantine layers. Red arrows indicate disease transmission. **τ_i_** is the time required for an infected person to become infectious and transmit the disease to the next layer. **b**, the daily cases as a function of time after reopening, if quarantine starts 7 days after infection (**τ_q_** = 7d). **c**, same as **b**, excerpt **τ_q_** = 11d. Error bars denote S.E.M.

In our model, 2-layer quarantine requires not only the confirmed carrier and its close contacts, but also their workplace, schools, homes, and communities to be quarantined. This additional layer leads to 950 +/− 69 quarantines per confirmed case. Nevertheless, at any given time, only a small percentage of the total population is under quarantine (Fig. S8).

In addition to 2-layer quarantine, schools, large businesses, and community events may need to be restricted if not enough cases are detected. If 70% of cases are detected, no restriction is necessary as long as 2-layer quarantine starts on day 8 (Figs. 1 and 4). If 60% of cases are detected, schools and businesses can reopen, but community events need to be restricted (Fig. 4). Community contacts outside schools, work, and home account for ~30% of daily contacts^12^ (Fig. S3).

**Figure 4.**
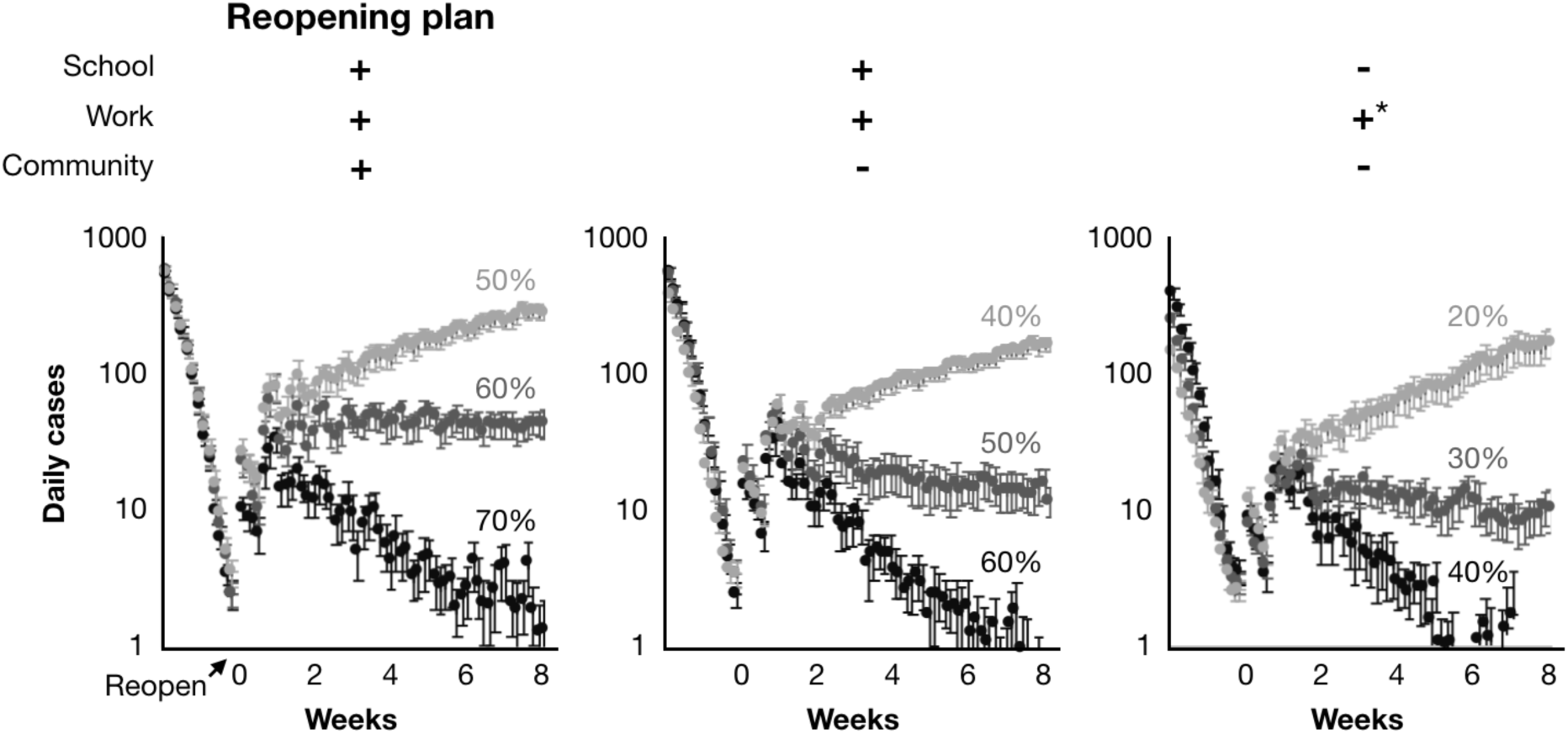
P_d_ determines whether a reopening plan will lead to resurgence. The daily cases as a function of time after reopening. Different colors correspond to different values of **P_d_**. * means only businesses with less than 2,000 employees are reopened. Error bars denote S.E.M.

If 30% of cases are detected, schools and large businesses (>2000 employees in our model) need to remain in virtual mode (Fig. 4). Due to their large size, schools and large businesses are hot spots for disease transmission. In our model, students are 66% +/− 12% more likely to transmit the disease than the rest of the population. Employees of large businesses, 94% +/− 5%. According to the model, had schools, large businesses, and community activities been shut down on March 3, the outbreak in Massachusetts would be limited to 3220 +/− 650 people and peaked by March 31 (Fig. S9).

Here we show the probability of resurgence for a given reopening open depends on P_d_, the percentage of cases detected, and τ_q_, the time quarantine starts (Fig. 1). This conclusion is based on the microsimulations of a hypothetical region similar to Massachusetts. To use the model for policymaking, however, it needs to be refined by local government records on population, employment, school, business, household, etc. The model can also be updated in the future as more data on disease transmission become available. The model can also be custom-built for regions other than Massachusetts. Nevertheless, for any given region, the more cases detected, the earlier quarantine starts, the less restrictive the reopening plan is expected to be.

The resurgence of infectious disease has significant impacts on public health and the economy. At the initial stage of reopening, it is safer to assume only a small percentage of cases (P_d_ = 30% or less) are detected late in the infectious window (τ_q_ = day 8 or later). In order to increase P_d_, it may be necessary to (i) test all symptomatic patients, (ii) test all close contacts of confirmed cases, and (iii) screen high-risk populations routinely. To decrease τ_q_, besides utilizing fast COVID-19 tests and contact tracking apps, it is also essential to consider the psychological and economic burden of testing and quarantine^13^. One possibility to offset such burdens is to issue stimulus checks to individuals and businesses affected by quarantines. 100,000 checks a day, $250 per check, cost $25million, which corresponds to 1.5% of daily GDP in Massachusetts^14^. 100,000 COVID-19 tests a day cost $5million^15^, or 0.3% of daily GDP. Therefore, encouraging more COVID-19 tests earlier is a small investment with a high return.

## Methods

### Disease transmission

In our microsimulation model, the disease spreads through three types of contacts. Contacts at school/work, home/residence, and community events account for 37%, 34%, and 29% of total contacts, respectively. This ratio is consistent with a recent study of social contacts in European countries^12^.

Each disease carrier makes up to four contacts at school/work every workday, three contacts at home/residence every night, and four contacts in a community event (see section region-specificity). The probability of infection upon contact is a free parameter in the model. We set it to 45%. This value is constrained by (i) the number of confirmed cases and (ii) the number of deaths reported by MDPH, and (iii) the number of conference attendees with COVID-19 symptoms reported by Biogen on March 3^16^.

### Detection and quarantine

Our model follows the timeline of the COVID-19 outbreak in Massachusetts. Once a case is detected, the disease carrier and its contacts are subject to quarantine. The number of detected cases between February 26 and April 9 is set by the number of confirmed reported daily by MDPH. Between April 10 and the reopen date (see section reopen), the number of detected cases is determined by min(2151, Pd*N). 2151 is the number of confirmed cases on April 9, Pd is the probability of detection, and N is the number of disease carriers between days 8 and 14 post-infection. After the reopen date, the number of detected cases is determined by Pd*N’, where N’ is the number of disease carriers on days **τ_q_** post-infection. We assumed that disease carriers under quarantine no longer go to work, school, community events, but continue to make contacts at home.

For 1-layer quarantine, contacts of the confirmed carrier between the day of infection and the day of detection are subject to quarantine. In comparison, 2-layer quarantine requires the school, workplace, home, and communities of the confirmed carrier and its contacts to be quarantined as well. In theory, it is possible to reduce the 2^nd^ layer to close contacts of the 1^st^ layer. However, this reduction would increase the workload of MDPH and might delay the start of quarantine. A slight delay might allow the virus to spread to the next layer and cause significant resurgence.

### Region-specificity

There are 9,000 daycares, 2,400 K-12 schools, and 100 colleges, with a total of 1.93 million students (see table 1). These numbers are estimated based on publicly available data on age distribution, student enrollment, and student/teacher ratio. They can be refined by local government records.

**Table 1.**
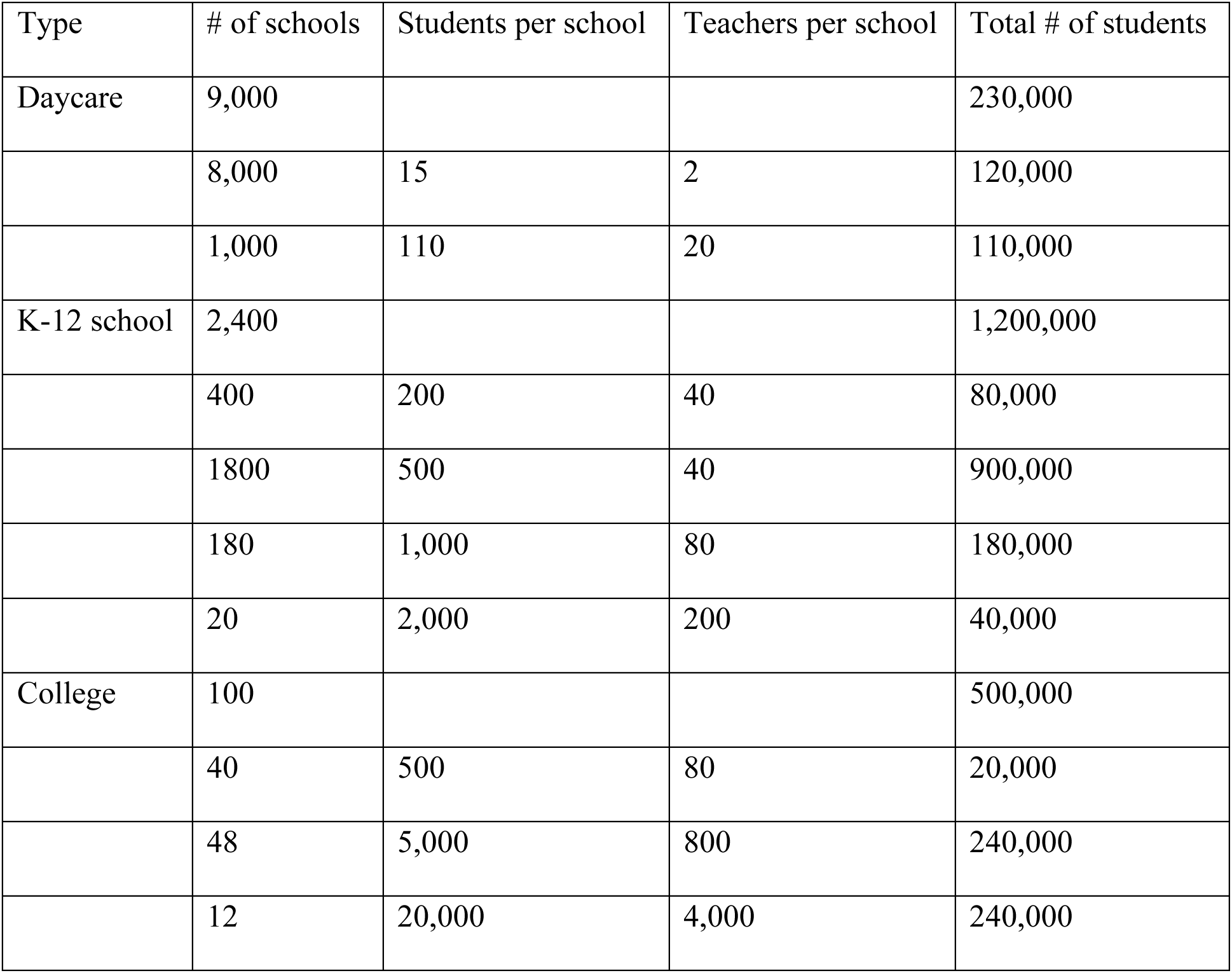
Daycares, schools, and colleges in the region-specific model

There are 197,715 employers with a total of 3.9 million employees (see table 2). Small (<19 employees), middle (20-99), large (100-499), and extra-large (500+) employers, account for 18%, 17%, 15%, and 50% of total workforce, respectively. This ratio is consistent with the national distribution of employer size^17^.

**Table 2.** Employers in the region-specific model

| Size | # of employers | Employees per employer | Total # of employees |
| --- | --- | --- | --- |
| 1-19 | 175,500 |  | 684,000 |
|  | 80,000 | 2 | 160,000 |
|  | 60,000 | 4 | 240,000 |
|  | 35,500 | 8 | 284,000 |
| 20-99 | 19,100 |  | 646,000 |
|  | 10,300 | 20 | 206,000 |
|  | 6,600 | 40 | 264,000 |
|  | 2,200 | 80 | 176,000 |
| 100-499 | 2,125 |  | 570,000 |
|  | 1,400 | 200 | 280,000 |
|  | 725 | 400 | 290,000 |
| 500+ | 990 |  | 1,900,000 |
|  | 500 | 800 | 400,000 |
|  | 270 | 2,000 | 540,000 |
|  | 200 | 4,000 | 800,000 |
|  | 20 | 8,000 | 160,000 |

There are 2.5 million households with 6.8 million residents (see table 3). Residents are randomly assigned to households. The household assignment is independent of the school/employer assignment.

**Table 3.** Households in the region-specific model

| # of households | # of adults per house | # of children per house | Total # of residents |
| --- | --- | --- | --- |
| 800,000 | 4 | 2 | 3,200,000 |
| 1,712,500 | 2 | 0 | 3,425,000 |
| 375 | 400 | 0 | 175,000 |
| 2,512,875 |  |  | 6,800,000 |

### Patient zero

We introduced patient zero in a large company (with 2,000 employees) on February 26, consistent with the COVID-19 outbreak in Massachusetts^16^. A conference organized by a Biogen between February 26 and 27 led to a dramatic increase of confirmed cases in the state^16^. We assumed that patient zero made eight contacts daily during the conference. That is two times the default number of contacts at work. This temporary doubling of contacts is needed to reproduce (i) an estimate of 50 conference attendees with flu-like symptoms on March 2 reported by Biogen^16^ and (ii) the dramatic increase of confirmed cases reported by MDPH the following week.

### Closure of school, business, and community

In our model, schools, community events, and businesses with more than 25 employees (except essential businesses) are closed on March 14. These closures are consistent with orders by Governor Baker and public announcements of schools, businesses, and event organizers. Upon closure, the disease can no longer transmit through these schools, businesses, and community events. In our model, all non-essential businesses are closed on March 22. Afterward, only 516,000 essential workers are allowed to work. In our model, the probability of infection is reduced by 50% on April 3. During that week, (i) CDC recommended wearing mask or face-covering; (ii) local grocery stores started to limit the number of customers inside the store; (iii) signs urging social distancing of 6 feet and redirecting foot traffic appeared in public places. We simulated the possible scenario that the probability of infection rebounds after reopening. If the probability is set to the default value of 45%, resurgence can still be prevented. For a given reopening plan, it requires more cases to be detected earlier.

### Validation of the model

In order to compare our model to the number of confirmed cases reported by MDPH, we assumed that cases are reported seven days after infection. This delay accounts for the incubation period (5-6 days^1,2^), consulting the primary care physician, ordering the test, and receiving the result. We assumed that confirmed deaths are reported 11 days after infection.

To calculate the serial interval, we analyzed all incidences, in which the disease is transmitted from a sender to a receiver. For each incidence, we calculated how long after the infection of the sender, the receiver is infected with the disease. We then averaged over all such incidences.

To calculate the reproduction number, we divided the number of infected people by the number of senders, before the closure of school, business, and community.

To quantify the likelihood of disease transmission by people affiliated with (i) schools, (ii) large businesses, and (iii) rest of the population, we calculated the number of senders in each population and then normalized by the size of the population.

### Reopen

In our model, reopening occurs when the total number of infected people does not change compared to the previous day. At the time of reopening, there are disease carriers who are infectious and yet to be detected.

## Data Availability

Available upon request

---

**Figure S1.**
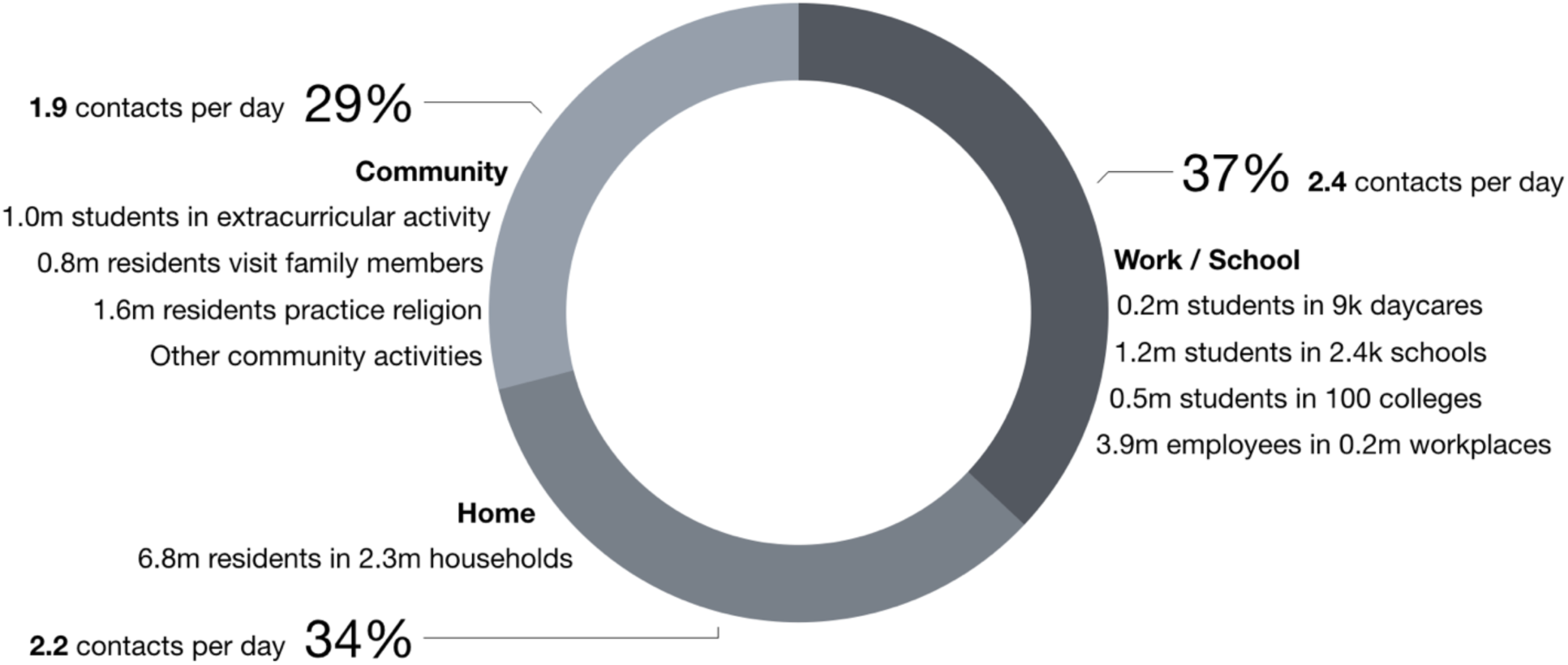
A microsimulation model applied to a region similar to Massachusetts. The percentage of close contacts in work/school, home, and community is consistent with a recent survey of daily contacts in European countries.

**Figure S2.**
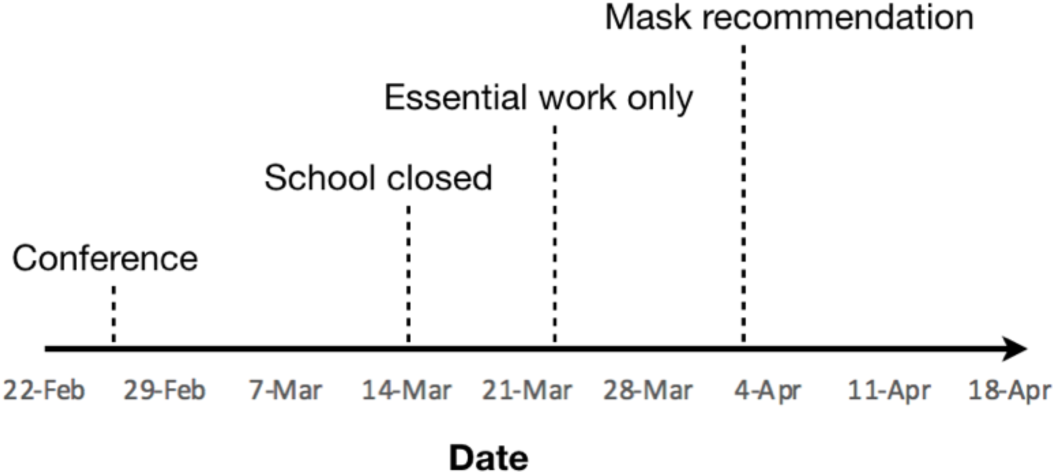
The timeline of COVID-19 responses in Massachusetts

**Figure S3.**
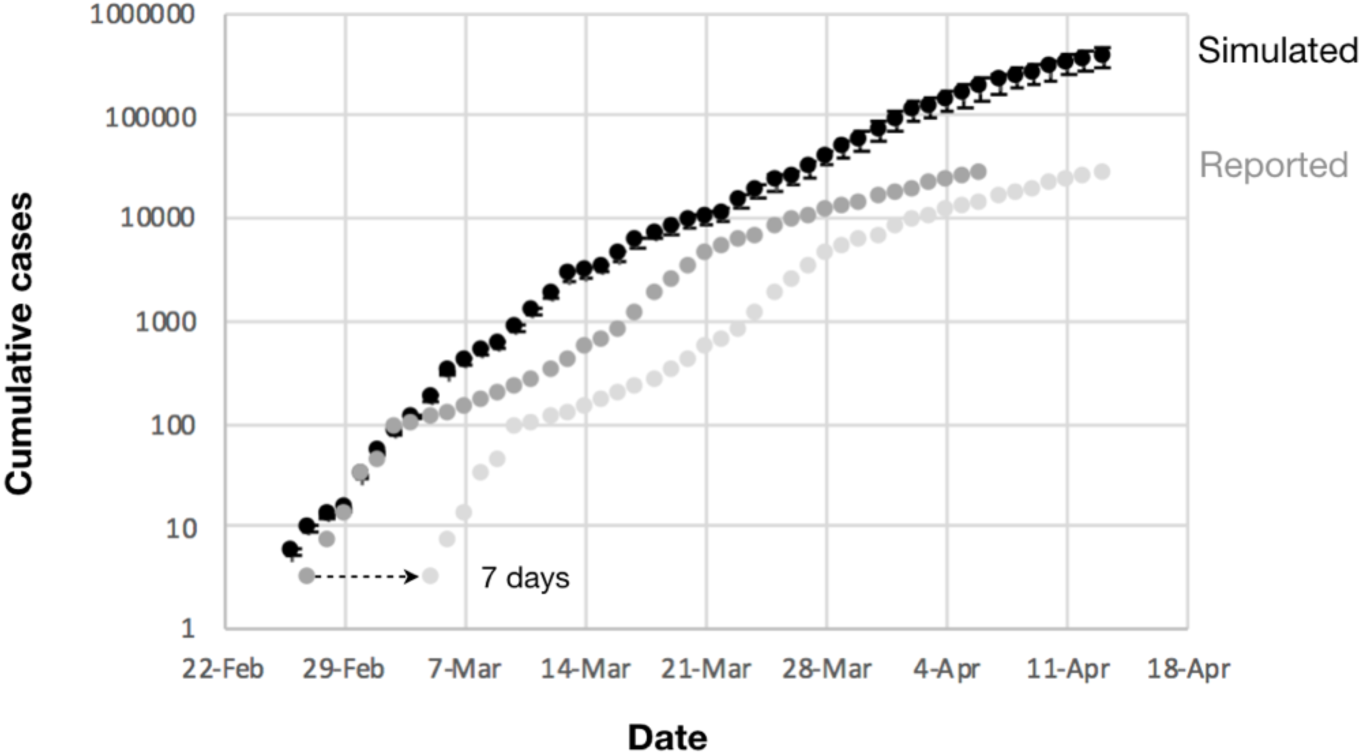
Cumulative cases as a function of time predicted by the simulation (black) and the number of COVID-19 cases reported by MDPH (light grey). Dark grey dots indicate the number of reported cases but shifted by seven days. Error bars denote S.E.M (n=20).

**Figure S4.**
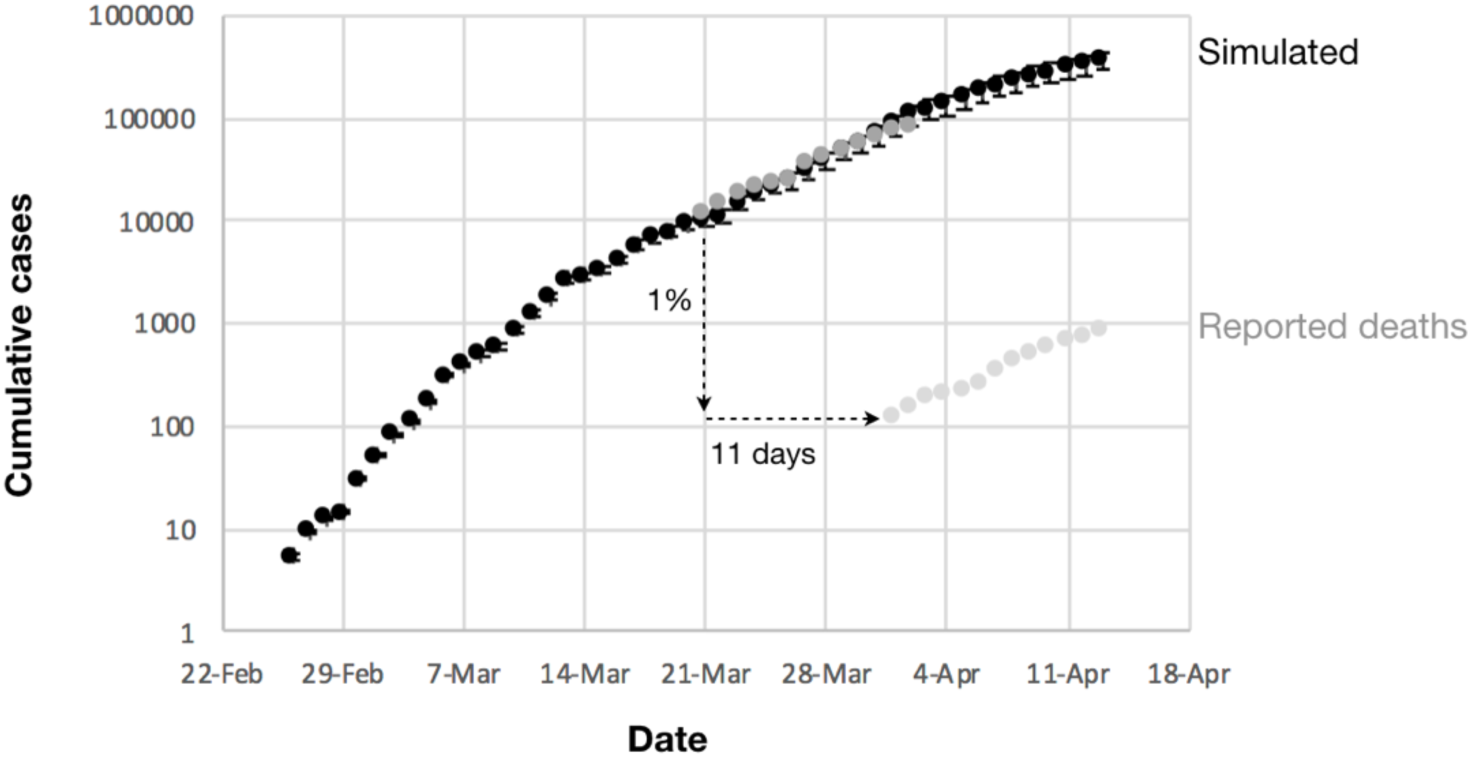
Cumulative cases as a function of time predicted by the simulation (black) and the number of COVID-19 related deaths reported by MDPH (light grey). Here the comparison is made on dates on which cumulative deaths are greater than 100. Dark grey dots indicate the number of reported deaths divided by an estimated death rate of 1% but shifted by 11 days. Error bars denote S.E.M (n=20).

**Figure S5.**
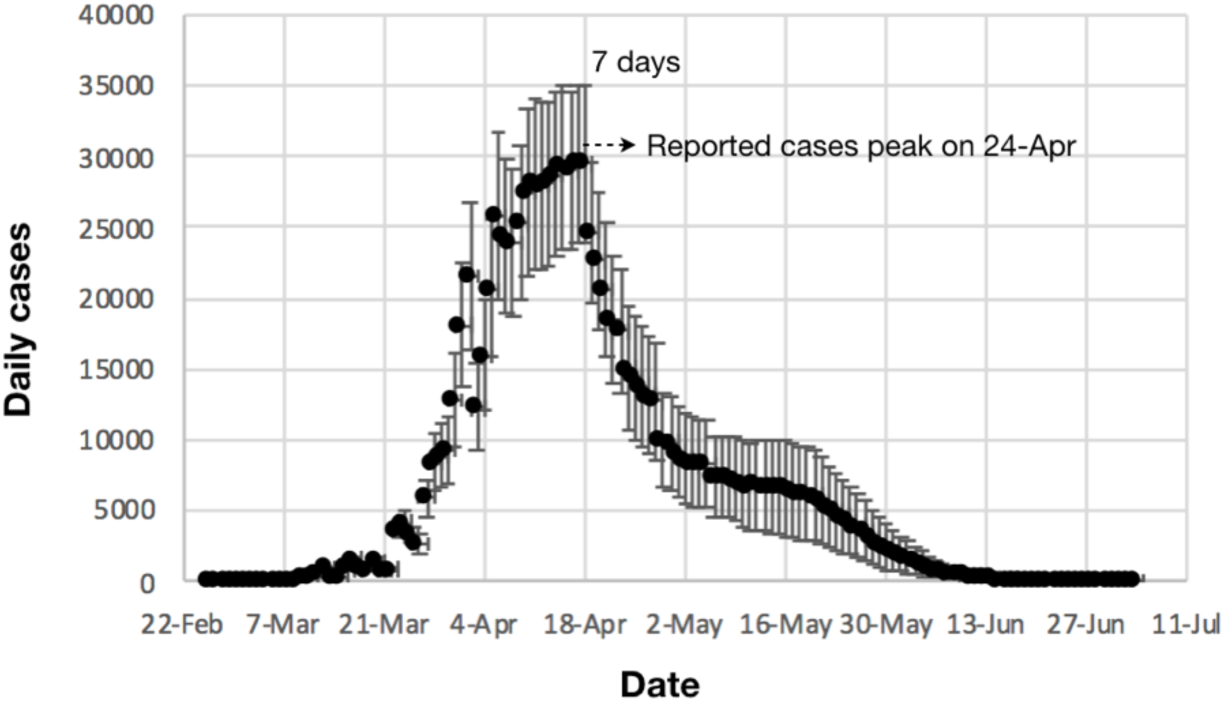
Daily cases as a function of time predicted by the simulation. The simulation predicts that the number of reported cases will peak on April 24, after an adjustment of 7 days to account the time between initial infection and test confirmation. Error bars denote S.E.M (n=20).

**Figure S6.**
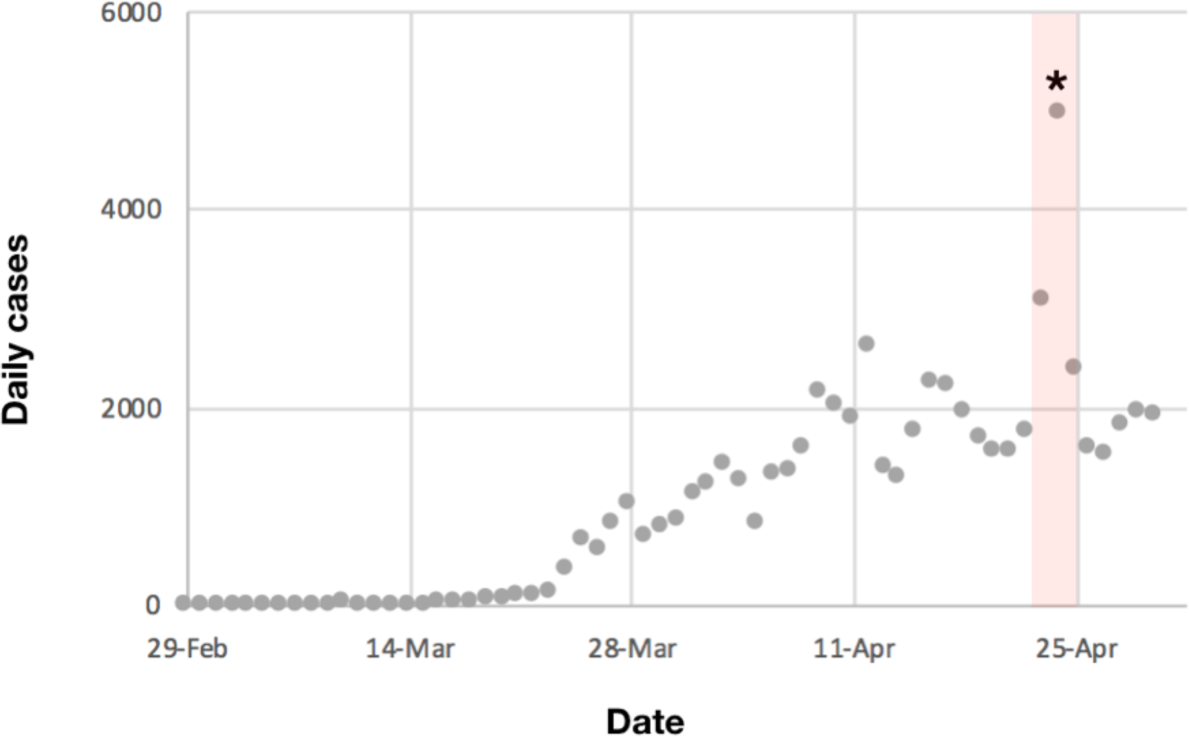
Daily cases as a function of time reported by MDPH. April 23-25 is highlighted in red. * denotes an adjustment, according to MDPH, due to previous reporting error.

**Figure S7.**
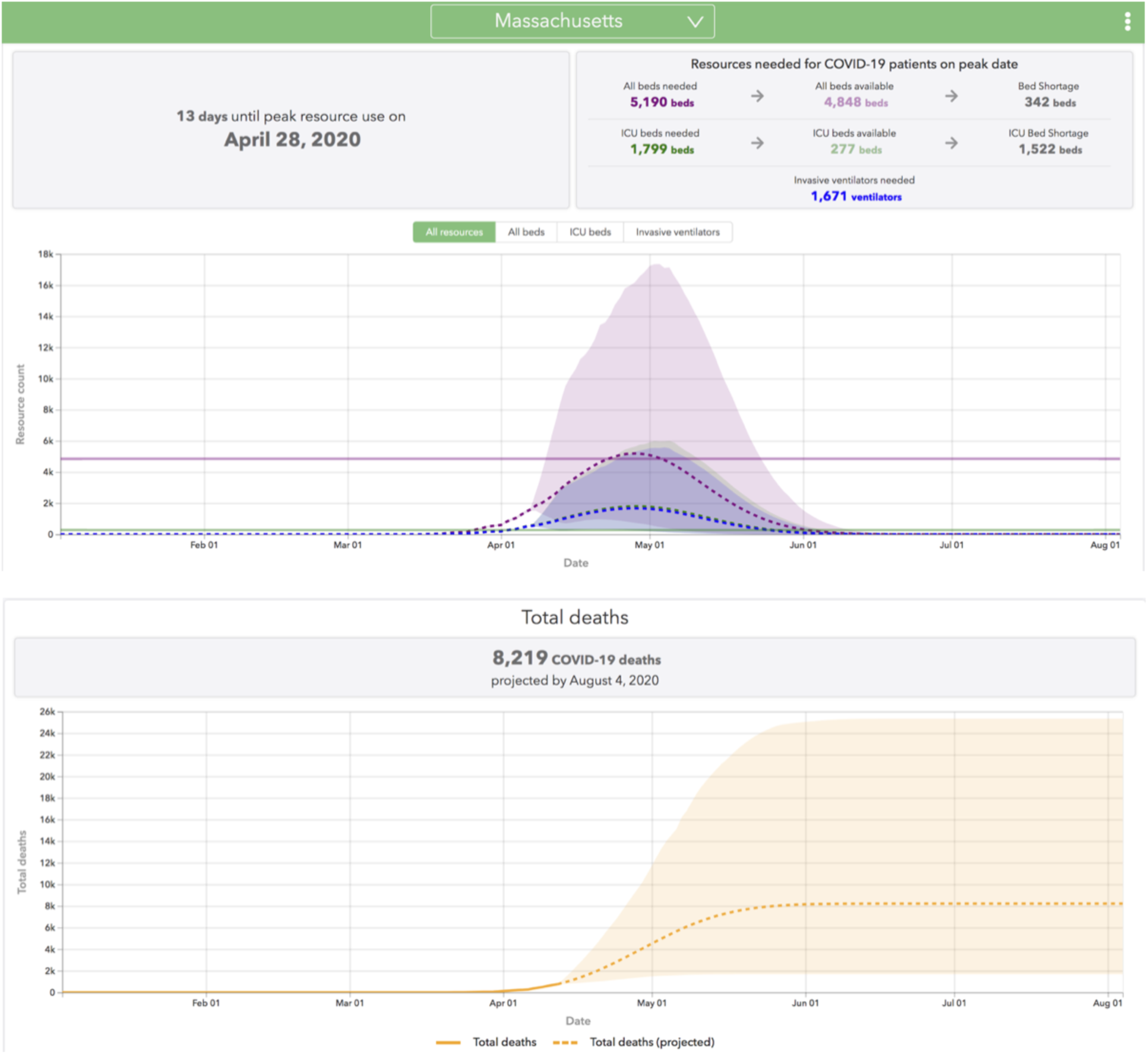
The University of Washington model’s prediction of medical resources needed and total deaths. The model predicts a peak date of April 28 for medical resources and 8,219 deaths by August 4.

**Figure S8.**
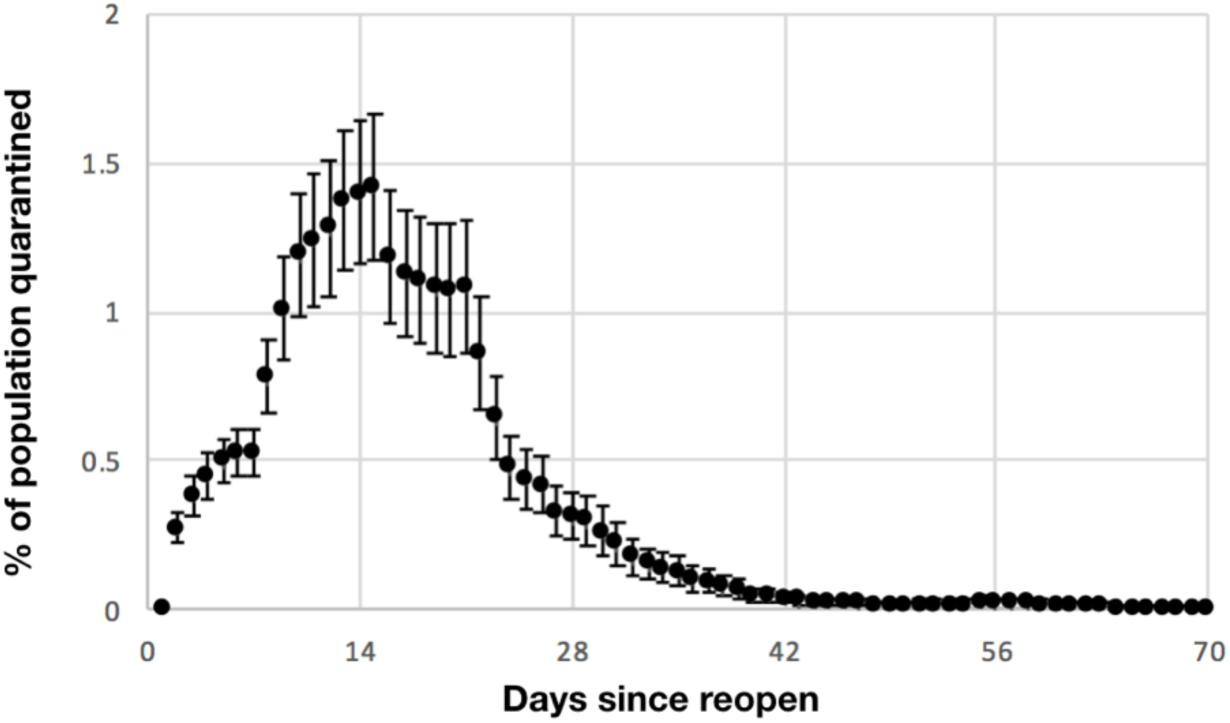
The percentage of the Massachusetts population under quarantine after reopening. Error bars denote S.E.M (n=20).

**Figure S9.**
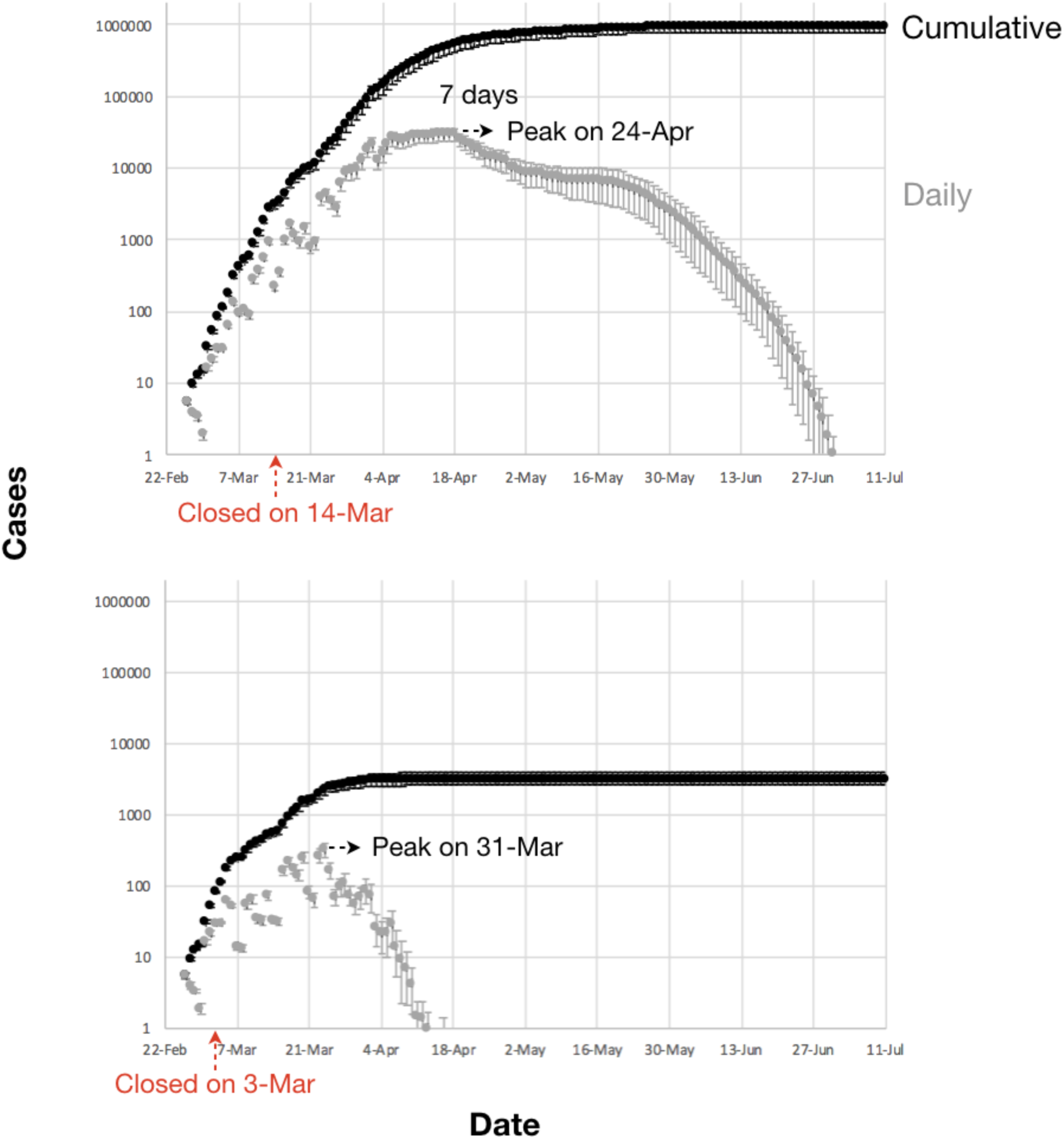
Cumulative and daily cases as a function of time in simulations in which schools, large businesses, and community activities shut down on March 14 (top) and March 3 (bottom). Error bars denote S.E.M (n=20).

## References

1 Anderson, R. M., Heesterbeek, H., Klinkenberg, D. & Hollingsworth, T. D. How will country-based mitigation measures influence the course of the COVID-19 epidemic? Lancet 395, 931–934, doi:10.1016/S0140-6736(20)30567-5 (2020).

2 Zou, L. et al. SARS-CoV-2 Viral Load in Upper Respiratory Specimens of Infected Patients. N Engl J Med 382, 1177–1179, doi:10.1056/NEJMc2001737 (2020).

3 CDC. Percentage of asymptomatic cases. (2020).

4 Enrico Lavezzo, E. F., Constanze Ciavarella, Gina Cuomo-Dannenburg, Luisa Barzon, Claudia Del Vecchio, Lucia Rossi, Riccardo Manganelli, Arianna Loregian, Nicolò Navarin, Davide Abate, Manuela Sciro, Stefano Merigliano, Ettore Decanale, Maria Cristina Vanuzzo, Francesca Saluzzo, Francesco Onelia, Monia Pacenti, Saverio Parisi, Giovanni Carretta, Daniele Donato, Luciano Flor, Silvia Cocchio, Giulia Masi, Alessandro Sperduti, Lorenzo Cattarino, Renato Salvador, Katy A.M. Gaythorpe, Imperial College London COVID-19 Response Team, Alessandra R Brazzale, Stefano Toppo, Marta Trevisan, Vincenzo Baldo, Christl A. Donnelly, Neil M. Ferguson, Ilaria Dorigatti, Andrea Crisanti. Suppression of COVID-19 outbreak in the municipality of Vo, Italy. Medrxiv (2020).

5 Halloran, M. E. et al. Modeling targeted layered containment of an influenza pandemic in the United States. Proc Natl Acad Sci U S A 105, 4639–4644, doi:10.1073/pnas.0706849105 (2008).

6 Team, I. C. C.-R. Impact of non-pharmaceutical interventions (NPIs) to reduce COVID-19 mortality and healthcare demand. (2020).

7 Andrew Ryan, K. L. a. S. D. Coronavirus deaths in Mass. are likely far higher than what’s been reported. Boston Globe (2020).

8 Zhang, Z. Prevent the resurgence of infectious disease with asymptomatic carriers. Medrxiv (2020).

9 Natsuko Imai, A. C., Ilaria Dorigatti, Marc Baguelin, Christl A. Donnelly, Steven Riley, Neil M. Ferguson. Report 3: Transmissibility of 2019-nCoV. (2020).

10 Donnelly, C. A. et al. Epidemiological determinants of spread of causal agent of severe acute respiratory syndrome in Hong Kong. Lancet 361, 1761–1766, doi:10.1016/S0140-6736(03)13410-1 (2003).

11 Lipsitch, M. et al. Transmission dynamics and control of severe acute respiratory syndrome. Science 300, 1966–1970, doi:10.1126/science.1086616 (2003).

12 Mossong, J. et al. Social contacts and mixing patterns relevant to the spread of infectious diseases. PLoS Med 5, e74, doi:10.1371/journal.pmed.0050074 (2008).

13 Coyle, B. Personal communication. (2020).

14 fred.stlouisfed.org. Total Gross Domestic Product for Massachusetts. (2020).

15 CMS.gov. Medicare Administrative Contractor (MAC) COVID-19 Test Pricing. (2020).

16 Andy Rosen, H. K., Kay Lazar, Jonathan Saltzman, Liz Kowalczyk, and Mark Arsenault. How the Biogen leadership conference in Boston spread the coronavirus. Boston Globe (2020).

17 Kiesrz, A. The impact of small business on the US economy in 2 extreme charts. Business Insider (2015).

